# Embedding the WHO’s Problem Management Plus (PM+) within Health and Integration Sectors in Switzerland

**DOI:** 10.1101/2025.08.19.25334041

**Authors:** Dharani Keyan, Julia Spaaij, Matthis Schick, Richard Bryant, Naser Morina

## Abstract

**Background:** The prevalence of common mental disorders in resettled refugee populations is nearly three times higher than in the general population. The World Health Organization (WHO) recommends *Problem Management Plus* (PM+), a brief non-specialist delivered intervention for reducing common mental disorders. To identify key factors to optimise implementation, we conducted a qualitative cross-sectional study to investigate contextual determinants shaping PM+ implementation during a nationwide scale-up of PM+ in Switzerland.

**Methods:** We conducted in-depth key informant interviews in the German-speaking and French-speaking parts of Switzerland (September 2023 and January 2025). Design and analysis of interviews was guided by the Consolidated Framework for Implementation Research. Themes were added both inductively and deductively and categorised into challenges and opportunities for wide-spread PM+ service delivery.

**Findings:** We interviewed 30 stakeholders: 12 policymakers, 5 government leaders, 5 specialists and implementers, and 8 non-specialist peer-providers. Whilst stakeholders were enthusiastic about the value of PM+, several limitations to its routine delivery were noted. These related to limited quality standards and control for non-specialist delivered interventions that prevented formal categorisation within the health system. Endorsement by professional associations including best-practice standards was thought to facilitate such formalised recognition. Prioritising mental health indicators within Cantonal integration programs was perceived to facilitate funding via the national integration agenda. All stakeholders pointed towards blended co-financing pathways to ensure to the longer-term sustainability of PM+ in Switzerland.

**Interpretation:** The routine delivery of PM+ within health and integration systems presented substantial challenges and opportunities. Longer term sustainability of PM+ may be feasible with blended financing, tailored Cantonal health and integration agendas, and established quality control standards for PM+ delivery that included formal recognition of non-specialists within the mental health system.

---

The refugee population has more than tripled around the world in the last decade ^1^. Following exposure to varied humanitarian crises arising from violence, armed conflict and natural disasters, many millions of people have sought refuge within other countries. Resettled refugees are substantially more vulnerable to mental disorders than the general population^2^. A recent meta-analysis indicated that the prevalence of common mental disorders (CMDs) including posttraumatic stress disorder (PTSD) and depression is nearly three times higher than that observed in the general population^3,4^. Evidence suggests that these mental health problems persist several years after resettlement ^5^. For host countries, this points to a clear need to adequately provide mental health services to meet the rising mental health needs of the population. This often entails planning for the provision of mental health services in ways that reorganizes available resources within existing systems of care. Importantly, the economic impacts of such planning have future implications for the integration of refugees and asylum seekers (RAS) beyond their initial period of resettlement within host nations.

Scalable psychological (or mental health) interventions have been developed to meet the needs of adversity-affected communities across both high-income countries (HICs), and low-and-middle income countries (LMICs)^6^. The World Health Organization’s Problem Management Plus (PM+) is one of the more widely tested scalable psychological interventions for the reduction of CMDs ^7^. Over five sessions, trained non-specialist providers deliver strategies in (a) stress management, (b) problem management, (c) behavioural activation, (d) strengthening of social supports, and (e) relapse prevention. PM+ has demonstrated its effectiveness in individual and group formats, including through remote video teleconferencing methods ^8–10^. A recent systematic review and meta-analysis of PM+ implementation found it to be effective in reducing distress and promoting positive mental health outcomes in varied populations exposed to distress across both LMICs and HICs^11^. For refugee populations, PM+ was found to reduce post-migration living difficulties in Switzerland ^12^, and reduced CMDs in the Netherlands ^13^.

Despite the enormous potential of PM+ to be embedded within existing care systems, its implementation within host country health systems has been slow. This is not surprising because the implementation of interventions, like PM+, requires consideration for a range of contextual determinants as it relates to service provision and planning across multiple levels – policy, institutional, organisational, and individual levels. These relate to barriers around the political will to scale up, professional apprehension of non-specialist delivered interventions for mental health care, lack of coordination between non-government organisations and national governments, lack of sustainable funding models, and apprehension about the use of task-sharing methods to treat CMDs^14–16^. There remains an information gap in relation to the static and dynamic contextual determinants shaping delivery of scalable mental health interventions. These factors relate to real-world indicators at multiple levels of host nation governments that may facilitate the routine uptake of scalable mental health interventions. We currently do not understand *under what conditions* and *by whom* existing resources can be reorganised (and/or mobilized) to accommodate the widespread implementation of PM+ within host country systems of care.

The current study is focussed on filling this gap by characterising key implementation findings gathered from Switzerland’s implementation of PM+. The Scaling-up Psychological Interventions in Refugees in Switzerland (SPIRIT) project is a national-scale-up study aiming to bridge the gap in mental health care for refugees by recruiting trained peer providers to deliver PM+. SPIRIT was launched following the success of its predecessor project, STRENGTHS, where PM+ was found to be both clinically- and cost effective in reducing distress and improving the wellbeing of Syrian refugees in varying countries in Europe and the Middle East^17^. Current knowledge of barriers and facilitators of PM+ implementation remains dominated by post-hoc application of frameworks and theories where there is a dearth of evidence derived from implementation science studies. To fill this gap, the current study investigated the static and dynamic determinants shaping widespread PM+ delivery in the German and French-speaking parts of Switzerland. We present the perceptions and experiences of key stakeholders from a health and integration systems perspective.

## Methods

### Study design

Study reporting adheres to the consolidated criteria for reporting qualitative research (COREQ)^18^. A cross-sectional qualitative study was conducted over 17 months following the launch of SPIRIT in 2021. SPIRIT is funded by the State Secretariet for Migration across all six asylum regions in Switzerland. Further information about the SPIRIT project is provided in the supplement. The Consolidated Framework for Implementation Research (CFIR) (version 1 and 2)^19,20^ was used to characterise and identify drivers or determinants of PM+ uptake. The CFIR, developed from a review of more than 500 implementation science frameworks^20^ examines multi-level dynamic and static factors influencing implementation of public health programs^21,22^. To the authors’ knowledge, no studies have used the CFIR to investigate a potential nation-wide scale-up of PM+ in LMIC or HIC settings. The following research questions were investigated from the perspectives of health and integration sector stakeholders: (1) challenges and opportunities for the introduction of PM+ within the health and/or integration systems; (2) perceived impacts of PM+ on refugee integration within Switzerland; and (3) the potential for nation-wide scale-up of PM+ for refugee communities in Switzerland. Data collection was undertaken during the mid-implementation phase of SPIRIT (September 2023 and January 2025), where PM+ implementation was underway across six asylum regions. This study was approved by the Ethics Committee of the Canton of Zurich (BASEC: 2023-00857).

### Study population and sampling

Study participants were identified across policy, institutional, organisational, and individual levels within health and integration sectors in Switzerland. Participant selection followed an ecological framework, assuming that future integration of PM+ into existing care systems will be shaped by multi-level factors. Both purposive and snow-ball sampling techniques were used. At the *individual* level, this included non-specialist providers delivering PM+ for RAS. At the *organisational management* level, it included professionals responsible for the training and coordination of non-specialist providers. At the *institutional* level, it included any professional, chief psychiatrist, or director within the Cantonal health and integration sector engaged with evidence-based mental health programmes, past or present. At the *policy* level, this included government officials or policy makers whose routine work comprised of decision-making on programmes for RAS within health and integration sectors.

Participants were approached by email, telephone, or in-person, where interviews took place either online or in-person, and this was determined based on their convenience and availability for participation. All invited stakeholders participated - there were no refusals. Participants provided informed oral consent prior to each interview. Data was obtained from six focus group discussions, and 16 key informant interviews: eight non-specialist providers, five organisational management-level stakeholders, five institutional stakeholders, and 12 policymaker and/or government personnel. Table 1 outlines stakeholder affiliations. To ensure confidentiality of data obtained, individual characteristics relating to age and gender have not been provided.

**Table 1.** Study participants interviewed by stakeholder group.

| Stakeholder group | Description | Number of interviews |
| --- | --- | --- |
| Federal health and integration leaders (i.e., policy makers) (n= 12) | National health leaders within the Federal Office of Public Health, the Federal Health Quality commission, State Secretariat of Migration, and Health Promotion Switzerland | 7 |
| Cantonal leaders (n = 5) | Chief Doctors, Lead psychiatrists of refugee and asylum seeker mental health services within central and western Switzerland asylum regions, Cantonal Commissioner for Health Promotion | 5 |
| SPIRIT Management team (n=5) | PM+ programme leader within a national NGO, Psychiatrists, Clinical psychologists, Coordinator of peer providers | 6 |
| Individual providers (n=8) | Non-specialist peer providers of PM+ | 12* |

\*three non-specialist providers were interviewed on two occasions.

### Procedures

The CFIR framework was systematically applied to guide design of interview guides, data collection, and evaluation. First, the *innovation* was defined as non-specialist delivered brief mental health interventions for RAS, with PM+ as the exemplar intervention. Second, the *inner setting* was defined as the Cantonal institutions (i.e., State institutions) coordinating PM+ implementation, with data collected from Zurich and Vaud - each represented a study site of the SPIRIT project. Third, the *outer setting* referred to the national Swiss context and implementation climate within which the *inner setting* operated. Accordingly, descriptions of the five CFIR domains were reviewed (intervention characteristics, outer setting, inner setting, characteristics of individuals, and process), and relevant constructs were mapped to SPIRIT as it related to scalable psychological intervention delivery. Two CFIR domains including *characteristics of individuals* and *implementation process* domains were deemed irrelevant as PM+ delivery was considered at a systems level (i.e., coordination through health, migration, integration sectors) instead of an individual and process level.

Initial topic guides for each stakeholder group were developed by DK and piloted via ongoing discussion with all study authors prior to formal data collection. To this end, the data collection strategy was iterative in that ongoing discussions between the first author (DK) and last author (NM) led to identification of new key informants. Additionally, interview topic guides were iteratively revised whereby debrief discussions after each stakeholder group (i.e., individual, and organisational management groups) led to adaptations of relevant topic guides for subsequent groups (i.e., institutional, and policy groups). Each interview collected information on participants’ professional background and experience, followed by open-ended questions.

In-depth interviews were conducted in English (n=11) and German (n =10), with a target duration of 60 minutes (range 45 – 120 minutes). Field notes and regular debrief discussions were held between study authors (DK, JS, NM, MS) to ensure that data interpretation was collaborative and driven by local needs. To verify the accuracy, stakeholder feedback was triangulated with supplementary data collection involving source documents relating to service provision for RAS in Switzerland (n = 7). These source documents were nominated by stakeholders and comprised of policy briefs and related government documents on RAS service provision and planning. This supplementary data collection ensured validity in reaching thematic saturation of CFIR constructs, and that no new related themes were identified. To validate extracted themes, preliminary findings were presented to stakeholders involved in the implementation SPIRIT project at dissemination meetings.

### Data management and analysis

All interviews were audio-recorded with informed consent (no refusals), pseudonymised, and transcribed via *Rev.ai*, an online Artificial Intelligence based tool (95% accuracy in English). German transcripts translated into English were checked for accuracy by JS and NM. Verified trasncriptions were transferred to *NVivo14* for coding and data analysis. Transcripts were first coded by thematic content analyses, guided by the CFIR framework in *NVivio*. Discussions were held between DK, JS, NM, and RB to agree on emerging themes. These were subsequently categorised by DK according to the CFIR constructs. Using an excel spreadsheet, each CFIR construct was categorised as a barrier, facilitator, or both in relation to PM+ service delivery for RAS in Switzerland. Source documents provided additional contextual understanding of stakeholder perspectives.

## Results

The final sample comprised of 30 in-depth interviews with 30 participants (12 national policy leaders, five institutional leaders, five organisational management team members, and eight peer providers; see Table 1). Findings are organised according to CFIR domains (see Table 2 and Figure 1).

**Table 2.** Summary of key challenges and opportunities to wide-spread PM+ implementation, by CFIR domain, as reported in in-depth stakeholder interviews and documentation review

| CFIR domains | Constructs | Challenges (data source) | Opportunities (data source) |
| --- | --- | --- | --- |
| 1. Intervention characteristics (i.e., PM+ as the exemplar scalable psychological intervention) | Relative advantage<br>Evidence base<br>Complexity<br>Adaptability<br>Design quality and packaging | • Need for localised evidence on the effectiveness of PM+ (IDI)<br>• PM+ is a novel service category not exclusively aligned with health, integration, or social initiatives. (IDI)<br>• Lack of formal titles or recognition of the helper creates confusion and undermines longer-term legitimacy of the role (IDI) | • Evidence of relative advantage in relation to available services for RAS (IDI)<br>• Establishing accreditation frameworks and quality control guidelines (e.g., minimum qualifications, supervision protocols, outcome tracking) could make PM+ viable within formal systems of care (e.g., health) (IDI) |
| 2. Outer setting (i.e., National Swiss context) | Local attitudes<br>Local conditions<br>Policies and Laws<br>Financing<br>Partnerships and connections | • Lack of national health policy on the role non-specialist/peer-delivered initiatives for mental health care (IDI, Doc)<br>• Decentralised governance of health leads to variation in initiatives for RAS across Cantons (IDI, Doc)<br>• Mental health screening is not mandated by law at the Federal asylum centres (IDI, Doc)<br>• Professional scepticism with public health (e.g., psychiatry) on the role of PM+ in adequately supporting RAS (IDI)<br>• No established financing pathway for non-specialist delivered interventions (Doc, IDI) | • Cantons can lead 'bottom-up' efforts to prioritise mental health and advocate for Federal alignment (e.g., budget support via SEM) (IDI)<br>• If screening measures were put in place at the Federal centres, and this was shared with Cantons, it could aid service-planning for longer-term integration support (IDI)<br>• Public health officials need education on the ways in which PM+ can be incorporated as a low-threshold initiative (e.g., collaborative care, stepped care pathways) (IDI)<br>• The FOPH mandate to promote Health equity offers justification for system-level support of PM+ (IDI, Doc)<br>• Establishing best-practice guidelines through professional association that included non-specialist delivered<br>interventions can help inform enforceable standards at the Federal level (IDI) |
|  |  |  | • Blended financing through State Secretariat of Migration, Health Promotion Switzerland, and Federal Health needs exploration (IDI, Doc) |
| 3. Inner Setting (i.e., Cantonal Institutions) | Compatibility<br>Mission alignment<br>Incentive Systems<br>Available resources<br>Tension for change<br>Implementation climate | • There is a lack of formal guidelines on where and how PM+ fits in within Cantonal health and integration systems (IDI)<br>• Equity in health service provision to RAS is not formally measured (Doc, IDI)<br>• Prioritisation of PM+ as a service for refugees varies across Cantons according to individual priorities (IDI) | • PM+ was perceived to align with other integration initiatives (e.g., job coaching, language lessons) in promoting Cantonal integration goals (IDI)<br>• Professional associations (e.g., psychiatry, clinical psychology) can advocate for PM+ with policy decision-makers (IDI)<br>• Development of specific health equity indicators for mental health care (e.g., access to interpreters) can create a pathway for PM+ accessibility (IDI) |

CFIR = consolidated framework for implementation research. SEM = state secretariat of migration. FOPH=Federal office of public health. PM+ = problem management plus. RAS = Refugees and asylum seekers. Data source were either documentation review (Doc) or in depth stakeholder interviews (IDI)

**Figure 1.**
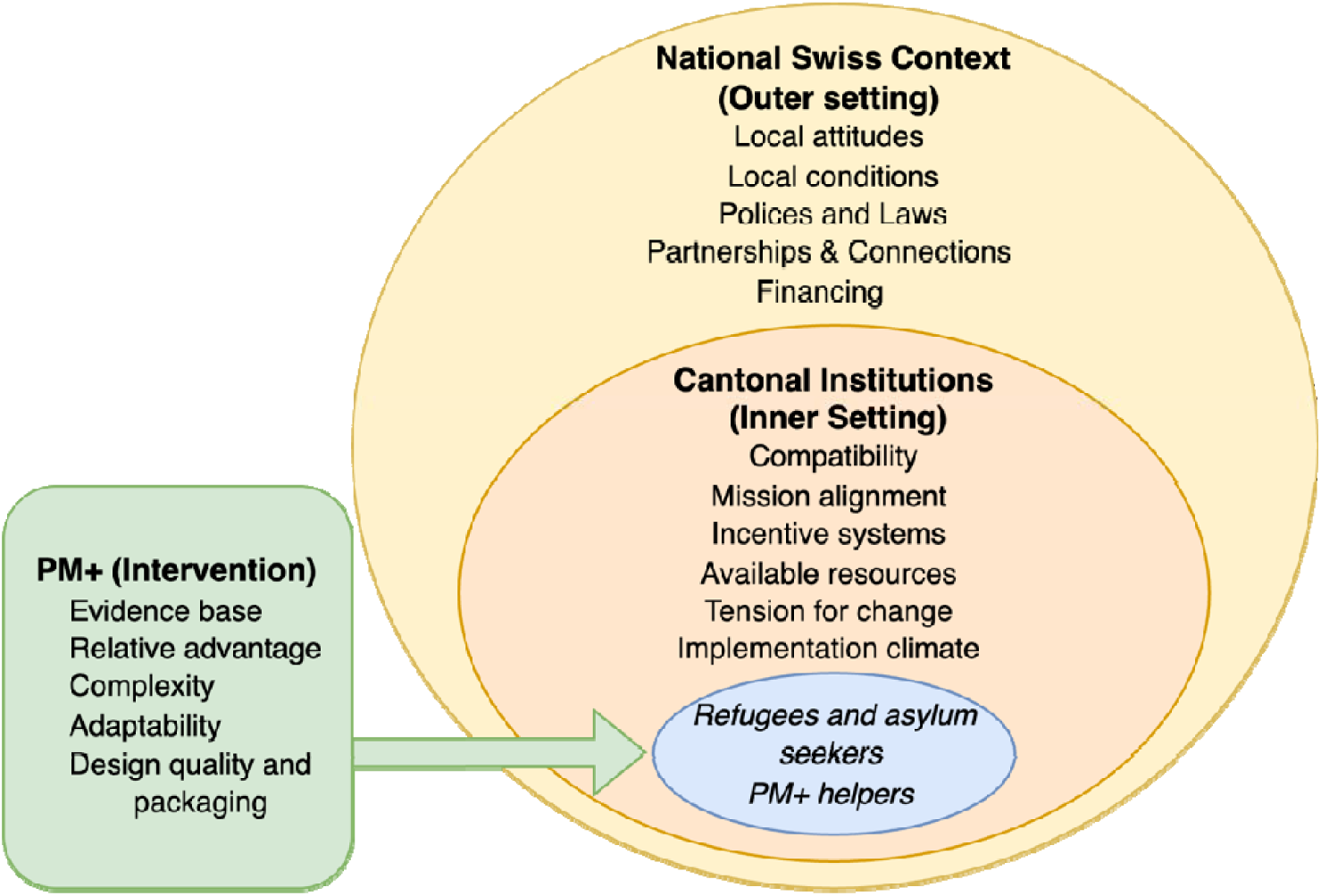
Examining determinants of PM+ implementation in Switzerland using the Consolidated Framework for Implementation Research.

### Characteristics of the PM+ intervention (CFIR domain one)

The following factors were characteristics of PM+ that influence its appeal as a (mental) health and/or integration initiative and in turn the likelihood of its successful future integration.

### Relative advantage

All stakeholders were enthusiastic about the potential of PM+ as a first line mental health care service for RAS, especially since such initiatives did not currently exist for refugees in Switzerland. NGO leaders, non-specialist providers, and Federal health leaders noted PM+ as having a marked advantage over utilising specialist services as a firstline intervention. These advantages included having peers speaking the same language as participants, and capacity to foster trust and authenticity. Federal health leaders viewed PM+ as an exemplar of ‘Patient and Public’ involvement in health care and viewed PM+ as serving as a positive first contact with the health system, potentially increasing future engagement with specialist care.

### Evidence base

Federal integration and health stakeholders stated localised evidence for the role of PM+ in preventing CMDs and influencing social and integration outcomes in Switzerland was insufficient. They expressed a desire to see the extent to which outcomes of SPIRIT supported these purported claims for PM+. Cantonal health leaders expressed an openness for peer-based delivery of PM+ and cited similar examples of this model of care that currently operated within other psychiatric services (i.e., substance misuse and addiction care). Health promotion stakeholders (see supplement for definition) similarly recognised use of peers as a first-line level of support and referred to parallel initiatives that currently existed for refugees (i.e., ‘Bridge builders’ program^23^).

### Complexity and Adaptability

Cantonal stakeholders noted that PM+ was perceived as a novel service category in that all stakeholders struggled to categorise it as an integration, health, social welfare initiative, or some combination of all. This lack of categorisation made it difficult to integrate PM+ within existing systems of care (i.e., health and/integration systems). For example, Cantonal health leaders expressed concern over siloing PM+ as just an integration initiative as this would likely limit where it could be delivered (e.g., such as primary health care settings). At the same time, it was unclear whether PM+ was preventative in that it could for example, reduce avoidable hospitalisation rates. The confusion over its role insofar as reducing CMDs, promoting social and/or economic integration was thought to hinder the buy-in from other specialist stakeholders including psychiatrists and physicians. For example, Cantonal psychiatry representatives noted that their respective colleagues were unclear whether PM+ was designed to replace specialist interventions for RAS. It was also unclear at what stage of the asylum process that PM+ should be prescribed to RAS.

### Design quality and packaging

Perceptions on how PM+ was packaged in terms its usability and design converged across stakeholders. First, ‘helpers’ (i.e., peer providers of PM+), as they are currently referred to within the SPIRIT project, expressed difficulties with explaining the nature of their role to beneficiaries and (mental) health specialists outside of the SPIRIT project (see Table 3 – intervention quotes). The lack of a formalised title for this role was perceived to somewhat undermine the acceptability of their role as a non-specialist provider of mental health care. Cantonal health leaders expressed the need for some form of certification (or accreditation) standards to be assigned to the role of a PM+ helper. To this end, peer providers expressed that the lack of accreditation pathways limited their professional identity. Policymakers expressed that the absence of standardised inclusion/exclusion criteria for recruitment of helpers would make quality assurance and financing more challenging in the future. To this end, they expressed a need for clear monitoring and evaluation standards for ongoing PM+ delivery. Health leaders and policymakers expressed that PM+ could be viable within existing systems of care (e.g., health system) if quality assurance and accreditation challenges are addressed (e.g., minimum qualifications, supervision protocols, outcome tracking). Federal health and Integration leaders expressed the potential for PM+ to be repositioned beyond the migrant and asylum context to other vulnerable groups (e.g., prisoners, health workers, schools), thus widening its relevance to policy makers.

**Table 3.**
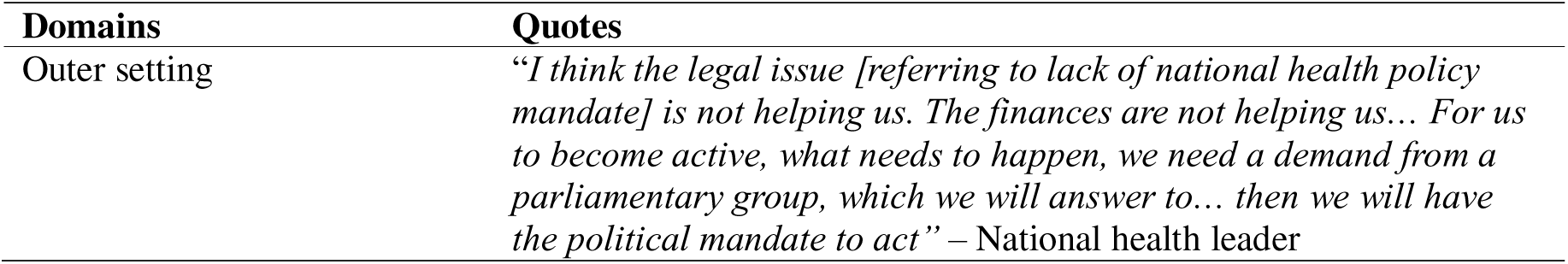
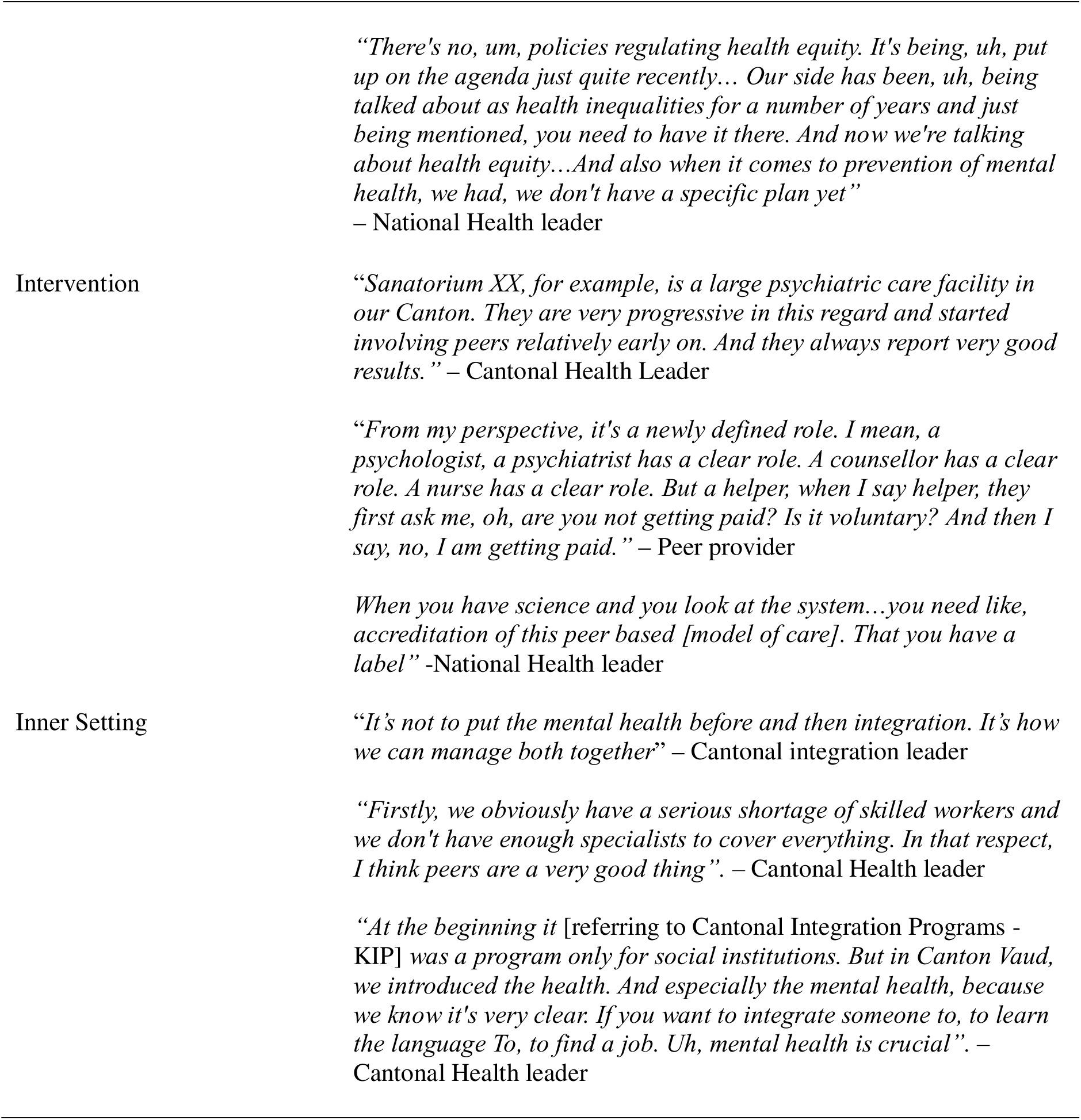
Interview Quotes organised by CFIR domains.

### National Swiss context (CFIR domain two: Outer setting)

The ‘outer setting’ domain summarises external factors shaping PM+ implementation, including local attitudes and conditions toward refugee needs at a national level, relevant policies and laws, and how funding guidelines are expected to impact PM+ service delivery.

### Local attitudes & Conditions

Swiss integration programs under the State Secretariat for Migration (SEM) prioritise practical integration (e.g., language, employment) with little attention to mental health. Decentralisation of health, where Cantons autonomously implement varied initiatives leads to inconsitences across different asylum regions. At the same time, Cantonal health leaders argued it as an opportunity for PM+ to be adopted in a tailored way, especially as RAS transition into Cantonal care. To this end, policymakers and Federal public health stakeholders advocated for sharing mental health screening outcomes with Cantons (following asylum procedures at Federal centers) to better align Cantonal service planning for long-term integration support. Health leaders and policy makers expressed that professional scepticism for PM+ rmeains, with some doubting that peer-delievered care sufficiently meets the mental health needs of RAS. Consequently, they expressed the importance of educating specialists (e.g., psychiatrists) and public health stakeholders on what “low-intensity” (or low-threshold) meant in practice, and how non-specialist peer-delivered care can operate within structured models (e.g., stepped care or collaborative care) without compromising the quality of care. To this end, Cantonal health representatives acknowledged workforce shotages and perceived PM+ as a potential ‘gap-filler’.

### Policies and Laws

Mental health screening is not mandated at Federal asylum centres. Federal integration leaders expressed concerns with introducing mental health screenings (e.g., for depression, PTSD) could complicate and potentially interfere with asylum proceedures at Federal centres. This discourages a centralised screening mandate and limits Federal level implementation of PM+. Cantonal health leaders argued that the lack of a national health policy that regulated how non-specialist (or peer-based) mental health care should be delivered, created uncertainty for PM+ integration within existing funding structures. Nevertheless, Federal quality commission (see supplement for definitions) stakeholders argued that the Federal Office of Public Health’s (FOPH) legal mandate to promote health equity offered a potential entry point for justifying system-level support for PM+ (see Table 3- outer setting quotes). To explore this path, they called for measurable (mental) health equity indicators, that currently did not exist.

### Partnerships and connections

Switzerland’s decentralised governance limits coordination across Federal and Cantonal institutions, and integration and health sectors. NGO, health and integration leaders argued that this limited cross-sector coordination of mental health services for RAS. For example, Cantonal health leaders stated that a routine referral pathway from Federal asylum centres to Cantonal mental health care systems did not exist. This was related to a lack of mental health screening at Federal centres. To this end, Federal and Cantonal stakeholders stated that Cantons could advocate for a centralised screening strategy or similar mandates as this served as a structural bridge between Federal and Cantonal systems.

### Financing

Stakeholders agreed that no established funding model exists for scalable, non- specialist delivered interventions. Current mandatory health insurance funds limited forms of specialist care (i.e., psychotherapy delivered by psychologists) for CMDs. The viability for blended financing by combining resources from health, integration, and prevention sectors was explored. The potential for the FOPH to support financing of PM+ through its health equity mandate was floated by Health Promotion stakeholders. Federal health and integration leaders argued that establishing best practice guidelines (e.g., through Swiss professional psychiatry associations) that included non-specialist delivered care can lead to enforceable standards under the health sector (e.g., FOPH), but this required partnerships between professional associations and Federal public health entities. Health leaders argued that co-financing PM+ under Cantonal Integration Programs (KIP)^24^ was an option given that Cantons defined integration priorities under the federal mandate of integration. To this end, the inclusion of mental health as part of their KIP outcomes, Canton Vaud had created a pathway for incentivisation of PM+ type services under the national integration agenda. Finally, funding through Health Promotion pathways was argued as a potential path if PM+ was found to be preventative in reducing the risk of CMDs. To this end, mandatory health insurance could be leveraged as partial funding for PM+ as funding for complementary medical services (e.g., physiotherapy) currently exist under this pathway.

### Cantonal settings (CFIR domain three: Inner setting)

The ‘inner setting’ examines the implementation climate and preparedness for PM+ service delivery within routine service provision pathways.

### Compatibility

Health and integration leaders perceived PM+ to fit within a tiered or stepped care model with the health system (e.g., prior to specialist support). Integration leaders perceived an alignment of PM+ with other integration services, where it had the potential to increase engagement with related supports (e.g., job coaching, language lessons). Canton Vaud Health representatives expressed a strong confidence in PM+ as a viable service for refugee populations, in the absence of clear alternatives. Cantonal integration leaders emphasised the need for positioning PM+ within a preventative frame as this was thought to increase its strategic fit within the integration agenda, as well as potentially aligning indicators with the FOPH mandate for promoting health equity.

### Mission alignment

All Cantonal leaders agreed that PM+ was not easily categorised as solely a health or integration initiative, and this created a challenge for institutional alignment and funding within Cantonal systems. Whilst PM+ conceptually aligned with the Federal health equity mandate, stakeholders were unclear on specific guidelines relating to where and how PM+ fits in. Federal health leaders emphasised a need for clinical leadership (e.g., professional psychiatry associations) to advocate for PM+ with policy decision-makers. To do this, it was recognised that educating clinical leadership on the proposed role of PM+ as complementary, rather than competing with specialist care, could ease their resistance and increase motivation for advocacy efforts. This was emphasised as an instrumental step in mobilising longer-term resource planning beyond project-based funding. For PM+ to be meaningfully integrated within Cantonal systems of care, health and integration leaders emphasised the need for routine mental health screening followed by immediate access to low-intensity interventions like PM+.

### Incentive systems & Available resources

Equity in health service provision to RAS is not measured (see table 3). Here, developing indicators of health equity (e.g., access to interpreters) will create an avenue to prioritise accessibility of PM+ to RAS. Health leaders argued that the lack of systematic funding for professional interpreters (e.g., Canton of Zurich) points to unequal access of (mental) health services. To this end, investing in professional interpreter services was viewed as a foundational step towards equitable (mental) health care and broader sustainability of PM+. Health leaders emphasised that indicators of health equity should be developed and prioritised by Cantons. For example, this could include linking access to interpreters for RAS with measurable health equity goals (e.g., reduced inequalities in access to health services).

### Tension for change and implementation climate

Health leaders noted that the priority for PM+ implementation varies across Cantons, and this is related to refugee population size across Cantons. To this end, mental health had yet to be embedded in many Cantonal integration agendas, and this currently limited opportunities for PM+ implementation. Canton Vaud was the first to institutionalise mental health as part of its Cantonal Integration Programs, and this offered a model for other asylum regions around the country. Canton of Zurich receives approx. 18% of RAS^25^, and this is leading to growing internal pressures to coordinate and invest in mental health initiatives. Health and integration leaders acknowledged an increasing awareness of the need to support the mental health and integration of RAS in parallel, and this likely creates an opening to align PM+ within their respective sectors (See Table 3 – inner setting). Here, NGO and integration leaders strongly viewed PM+ as a critical intervention for RAS given a lack of alternatives. At the same time, formal recognition of non-specialist delivered interventions by Federal and/or Cantonal health directorates was argued to be essential for long-term sustainability. For health directorate endorsement (i.e., FOPH), policy makers emphasised the need for clear operational standards for PM+ implementation (i.e., quality assurance standards) and that this was a major barrier to securing sustainable funding in the longer-term.

## Discussion

The current study provides key insights into the challenges and opportunities for systematically implementing PM+ through health and integration systems in Switzerland. To our knowledge, this is the first study to apply the CFIR^19,20^ to understand system-level barriers and enablers to PM+ implementation, especially within a decentralised host nation setting. Many of the challenges identified have been previously reported, such as the professional scepticism of scalable psychological interventions, and lack of sustainable funding pathways ^14,15^. The current study reveals important unanticipated considerations for widespread implementation of PM+ within health and integration systems in Switzerland. The proposed strategies within current policy frameworks highlight how current resources may be mobilised to meet the high need for evidence-based scalable psychological interventions for RAS.

Framed through the intervention characteristics domain, all leaders, providers, and policy decision makers readily acknowledged the *relative advantage* of PM+ as an important first-line intervention to support refugee mental health and their integration within Cantons. At the same time, the need for an *evidence base* that was localised to Switzerland insofar as the potential preventative and/or therapeutic role of PM+ in facilitating refugee integration was highlighted by policy decision makers. Despite the potential for PM+ as a low-threshold mental health initiative, several Cantonal health leaders described a hesitation from professional institutions to formally endorse this type of intervention. This related to the perceived unclear objective of PM+ in terms of whether it competed with specialist care services. To this end, it was acknowledged that formal mental health associations and leading psychiatry representatives required education on how PM+ could be embedded within stepped care and/or collaborative care initiatives for RAS. This importantly highlighted the need for developing best-practice recommendations when choosing PM+ vs. specialist care as a first-line service for this population in Switzerland. Integration and Health leaders put forth a need for strategic positioning of PM+ as a low threshold preventative service that complemented, rather than replaced specialist mental health care.

The most concrete barrier to systematic PM+ implementation in Switzerland were structural characteristics of the Swiss setting involving its siloed governance of health, welfare, and integration systems as it pertained to refugee and asylum seeker care within Cantons. Specifically, the lack of a unified mental health strategy for RAS arriving in the country impacts the extent to which Federal health entities (e.g., FOPH) can legally enforce coordination of mental health services for RAS within Cantonal institutions. Here, the *relative priority* and urgency for mental health support was thought to vary across the 26 Cantons, and this was likely shaped by refugee intake and perceived *tension for change* in meeting the unmet needs of the population. One Canton provided an exemplar of high implementation readiness where it had advocated for mental health support as part of its individualised Cantonal Integration Program (KIP). In turn, this had opened a co-financing pathway through existing integration programs. Further, PM+ was perceived as a novel service category (i.e., *design quality and packaging*) and this in part was due to the lack of formalised recognition of the role of PM+ ‘helpers’, and limited formal guidance on use of non-specialist interventions for refugee mental health care. This importantly made service planning within existing care pathways of health and/or integration a challenging pursuit. Consistent with past evidence^15,16^, stakeholders reiterated that PM+ did not readily fit within mandatory health insurance service provisions. To this end, the absence of quality control standards for PM+ service delivery was a considerable barrier for its resource allocation within mandatory health insurance.

Given the lack of a clear financing pathway for non-specialist delivered (mental) health interventions for refugees, the proposal for co-financing was a prominent theme in the *financing* and *partnerships and connections* constructs. Within an integration systems perspective, the *compatibility* of PM+ with other integration initiatives available to RAS including job coaching, language lessons, and social support was perceived to be high. Integration leaders argued that this provided a rationale for its fit and co-financing within the broader integration agenda. To this end, there was agreement on the need for mental health screening to be mandated (at Federal centres or upon Canton entry) that was in turn linked to referral pathways to PM+ service delivery. Developing incentives in the form of linking PM+ delivery to measurable integration goals (e.g., social, and economic integration) was recommended. Despite the relative barriers to including PM+ within the formal health system, policy makers and health leaders emphasised that principles of health equity provided a rationale and pathway for co-financing of PM+ as part of the health system. To this end, if Federal entities incentivised Cantons to meet their health equity goals (e.g., reduce inequalities in access to mental health care by refugee populations), the need for interpreters could be championed and in turn this paved a path for longer-term sustainability of PM+ within the health system. However, it was proposed that national best-practice guidelines and accompanying quality standards (e.g., supervision, evaluation, accreditation guidelines) needed to be developed to facilitate the process of formal funding for PM+ through mandatory health insurance. Finally, mandating centralised mental screening of RAS was advocated by Cantonal health representatives as a prerequisite for PM+ referral pathways. Supporting this proposition are recommendations from the 2019-2020 annual report by the National Commission for the Prevention or Torture that recommends early identification of vulnerable persons within Federal and Cantonal reception centres^26^.

Our study had several limitations. We intentionally chose to investigate implementation within the German- and French speaking parts of Switzerland, where stakeholders had expressed active motivation in sustaining PM+ implementation beyond the SPIRIT project. Although the German- and French speaking parts represent a much higher proportion of the refugee population, we acknowledge that the representativeness is limited, especially where perceptions of support for RAS may differ, and where the proportions of refugee intake vary across other Cantons. As such, we caution broad generalisability of our findings. All interviews were conducted within official settings (either in-person, or online), that had no relationship to ongoing SPIRIT project activities. Despite this, all stakeholders were generally positive towards PM+ implementation and this in part may reflect acquiescence in official settings. At the same time, we made attempts to limit such social desirability biases by presenting participants with written information relating to how anonymity of data will be maintained. Finally, stakeholder interview guides were not designed to address all CFIR constructs. As such, missing constructs from our findings reflects our study protocols rather than an indication of the importance of these constructs in future studies of systems perspectives on PM+ implementation.

To our knowledge, this is the first study to systematically apply the CFIR to design, conduct and analyse challenges and opportunities for widespread PM+ implementation within any settings. The current findings validate previous qualitative studies on barriers towards widespread implementation of non-specialist delivered interventions for CMDs. Our study additionally applies a health and integrations systems lens in evaluating the likelihood of its successful uptake in a high-income host nation. Importantly, policy maker, institutional and health leader perspectives on likely context-specific determinants of PM+ implementation in a high-income setting have previously not been uncovered. Together, findings provide a necessary foundation for developing targeted implementation strategies for widespread PM+ implementation in Switzerland.

## Contributors

DK, JS, MS, RB, and NM conceptualised the research questions and study design. DK, and JS contributed to data acquisition. DK, NM and JS accessed and verified the data. DK did the initial coding and JS carried out coding verification. DK and JS coded the data for this analysis. DK drafted the manuscript. All authors had full access to the data, and contributed equally to manuscript review and editing and approved the final version.

## Data sharing

Our full study protocol can be made available upon reasonable request. The interview transcripts analyzed during this study are not available because these files are personally identifiable. The data files do not contain names; however, files would be identifiable to a person familiar with the Swiss health and integration sectors given that study participants were identified by their specific roles and responsibilities within these care systems.

## Acknowledgements

Funding for SPIRIT is provided through Health Promotion Switzerland, whereas the single Cantonal implementing partners are financed through the State Secretariat of Migration, and respective Cantonal authorities. We are very grateful to the study participants who took part in the interviews. The findings and conclusions in this paper are those of the authors and do not represent the official position of Federal and Cantonal Institutions.

